# Significant reduction in humoral immunity among healthcare workers and nursing home residents 6 months after COVID-19 BNT162b2 mRNA vaccination

**DOI:** 10.1101/2021.08.15.21262067

**Authors:** David H. Canaday, Oladayo A. Oyebanji, Debbie Keresztesy, Michael Payne, Dennis Wilk, Lenore Carias, Htin Aung, Kerri St. Denis, Evan C. Lam, Christopher F. Rowley, Sarah D. Berry, Cheryl M. Cameron, Mark J. Cameron, Brigid Wilson, Alejandro B. Balazs, Christopher L. King, Stefan Gravenstein

## Abstract

High COVID-19 mortality among nursing home (NH) residents led to their prioritization for SARS-CoV-2 vaccination; most NH residents received BNT162b2 mRNA vaccination under the Emergency Use Authorization due to first to market and its availability. With NH residents’ poor initial vaccine response, the rise of NH breakthrough infections and outbreaks, characterization of the durability of immunity to inform public health policy on the need for boosting is needed. We report on humoral immunity from 2 weeks to 6-months post-vaccination in 120 NH residents and 92 ambulatory healthcare worker controls with and without pre-vaccination SARS-CoV-2 infection. Anti-spike and anti-receptor binding domain (RBD) IgG, and serum neutralization titers, were assessed using a bead-based ELISA method and pseudovirus neutralization assay. Anti-spike, anti-RBD and neutralization levels dropped more than 84% over 6 months’ time in all groups irrespective of prior SARS-CoV-2 infection. At 6 months post-vaccine, 70% of the infection-naive NH residents had neutralization titers at or below the lower limit of detection compared to 16% at 2 weeks after full vaccination. These data demonstrate a significant reduction in levels of antibody in all groups. In particular, those infection-naive NH residents had lower initial post-vaccination humoral immunity immediately and exhibited the greatest declines 6 months later. Healthcare workers, given their younger age and relative good-health, achieved higher initial antibody levels and better maintained them, yet also experienced significant declines in humoral immunity. Based on the rapid spread of the delta variant and reports of vaccine breakthrough in NH and among younger community populations, boosting NH residents may be warranted.

## Research Letter

The very high overall COVID-19 morbidity and mortality among nursing home (NH) residents led to their prioritization for early vaccination. In the US, most NH residents received BNT162b2 mRNA vaccination both because of its earlier Emergency Use Authorization approval and availability. NH residents that never had SARS-CoV-2 infection before the time of the vaccination series produced ∼¼ the antibody to this vaccine than that produced by healthy younger individuals.^1,2^ With the rise of breakthrough infections in NH residents and others with immunocompromise, there is a significant need for characterization of the durability of immunity to inform public health policy. We report on 2-week and 6-month post-vaccination antibody levels from 120 NH residents (aged 48-100 years, median 76) and 91 ambulatory healthcare worker controls (aged 26-78 years, median 48) with and without SARS-CoV-2 infection prior to vaccination. We assessed anti-spike and anti-receptor binding domain (RBD) IgG, and serum neutralization titers, using a bead-based ELISA method and pseudovirus neutralization assay.^1^ Anti-spike, anti-RBD and neutralization geometric mean levels dropped more than 84% over 6 months’ time in all groups (Fig. 1a-c) irrespective of prior SARS-CoV-2 infection. At 6 months post-vaccine, 70% of the infection-naive NH residents had neutralization titers at or below the lower limit of detection (LLD, 1:12 titer) compared to 16% at the LLD at 2 weeks after full vaccination (Supplemental Table 1). Controls and NH residents who survived prior infection had 19% and 35% respectively at LLD at 6 months post-vaccination. These data demonstrate significant 6-month antibody decline in all groups.

**Figure 1.**
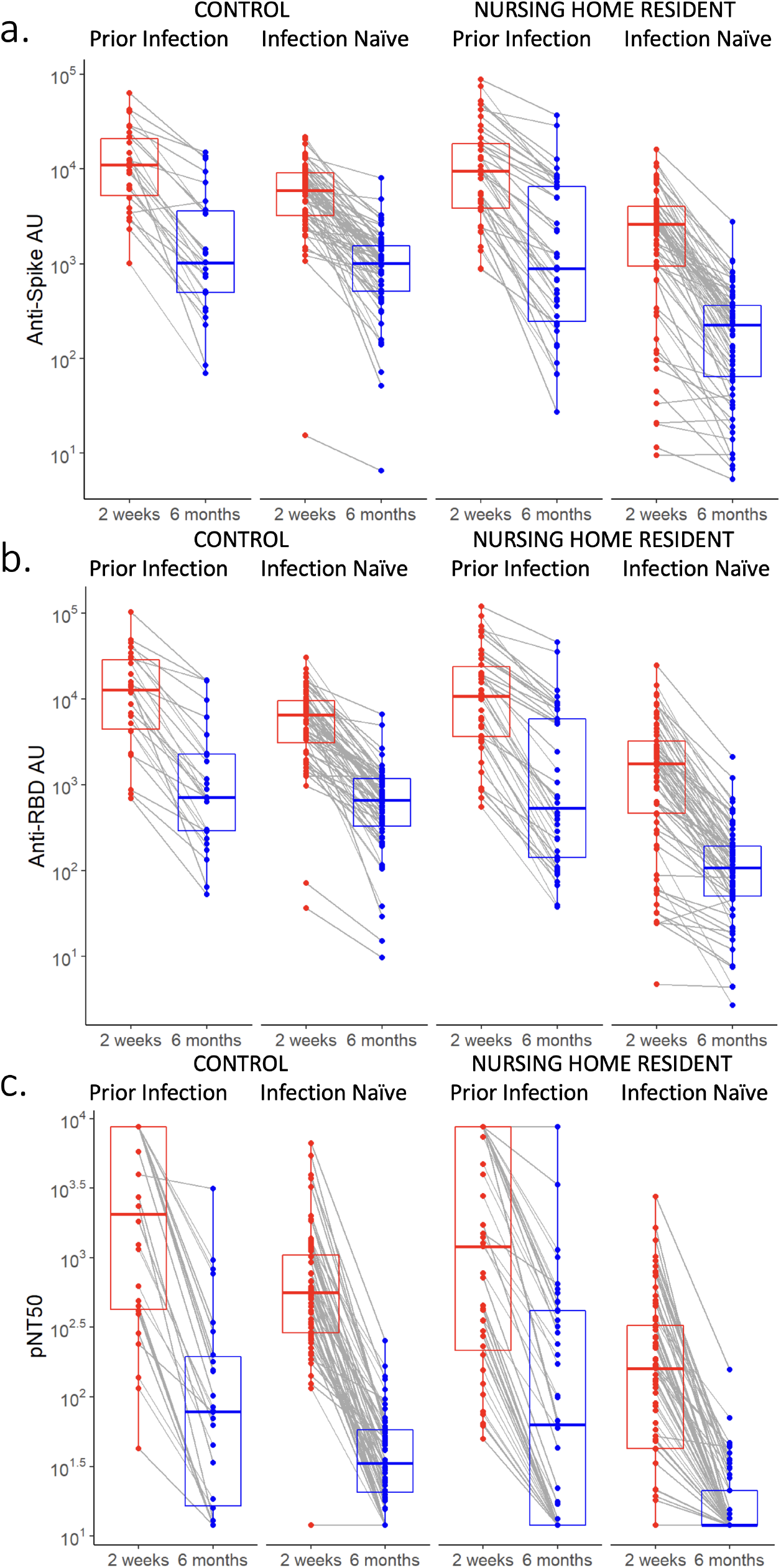
Antibody levels 2 weeks and 6 months after BNT162b2 mRNA vaccination in healthcare workers and nursing home residents, with and without SARS-CoV-2 infection prior to vaccination. Post-vaccination anti-spike arbitrary units (AU) (Figure 1a.), anti-receptor binding domain (Figure 1b), and pseudovirus neutralization (pNT50) (Figure 1c) are shown. Residents refers to NH residents and Control refers to younger healthcare workers. Prior infection refers to antibody levels at the given time points in individuals vaccinated after recovering from earlier SARS-CoV-2 infection, and infection naive refers to individuals vaccinated without prior SARS-CoV-2 infection. Geometric means in each group were assessed using paired t-tests of log-transformed values among subjects with both measures present. P values comparing the difference between 2 weeks and 6 months in all clinical groups were p<0.001. Subjects with large increases (>100%) of anti-spike, anti-RBD, or neutralizing titers from 2 weeks to 6 months were excluded due to presumed SARS-CoV-2 infection after vaccination.

An improved understanding of the clinical consequences of this drop in humoral immunity is urgently needed to optimally inform boosting strategies and policy that are being actively considered in the recent CDC recommendation. However, in the absence of clinical evidence, extrapolating from laboratory values may be necessary. NH residents and healthcare workers were amongst the earliest populations vaccinated in the US and elsewhere, resulting in the longest time for immunity to wane. In particular, those infection-naive NH residents had lower initial post-vaccination humoral immunity immediately and exhibited the greatest declines 6 months later. Healthcare workers, given their younger age and relative good-health, achieved higher initial antibody levels and better maintained them, yet also experienced significant declines in humoral immunity. The rapid delta variant spread, vaccine breakthrough in NHs and community,^3-5^ and rapid antibody decline support CDC’s recommended boosting NH residents to curb spread or prevent severe illness.

**Supplemental Table 1.** Proportion at the lower limit of detection (LLD, 1:12 titer) with pseudovirus neutralization assay (pNT50).

|  | 2 weeks<br>post-vaccination | Fisher's exact test<br>*p*-value | 6 months<br>post-vaccination | Fisher's exact test<br>*p*-value |
| --- | --- | --- | --- | --- |
| **Naïve** |  |  |  |  |
| Control | 1/64 (2%) | 0.005 | 10/64 (16%) | <0.001 |
| NH resident | 11/73 (16%) |  | 51/73 (70%) |  |
| **Prior infection** |  |  |  |  |
| Control | 0/26 | NS | 5/26 (19%) | 0.19 |
| NH resident | 0/43 |  | 15/43 (35%) |  |

## Data Availability

We will evaluate collaboration to provide access to the data.

